# The Novel Coronavirus Disease (COVID-19): A PRISMA Systematic Review and Meta-analysis of Clinical and Paraclinical characteristics

**DOI:** 10.1101/2020.04.19.20071456

**Authors:** Hamidreza Hasani, Shayan Mardi, Sareh Shakerian, Nooshin Taherzadeh-Ghahfarokhi, Parham Mardi

**Author notes:** Corresponding Author:* Parham Mardi, MD, Student Research Committee, Alborz University of Medical Sciences, Karaj, Iran.

## Abstract

An outbreak of pneumonia, caused by a novel coronavirus (COVID-19) was Identified in China in Dec 2019. This virus expanded worldwide, causing global concern. Clinical, laboratory and imaging features of this infection are characterized in some observational studies. We undertook a systematic review and meta-analysis to assess the frequency of clinical, laboratory, and CT features in COVID-19 patients.

We did a systematic review and meta-analysis using three databases to identify clinical, laboratory, and CT features of rRT-PCR confirmed cases of COVID-19. Data for 3420 patients from 30 observational studies were included.

Overall, the results showed that fever (84.2%, 95%CI 82.6-85.7), cough (62%, 95%CI 60-64), and fatigue (39.4%, 95%CI 37.2-41.6%) were the most prevalent symptoms in COVID-19 patients. Increased CRP level, decreased lymphocyte count, and increased D-dimer level were the most common laboratory findings. Among COVID-19 patients, 92% had a positive CT finding, most prevalently GGO (60%, 95%CI 58-62) and peripheral distribution (64%, 95%CI 60-69).

These results demonstrate the clinical, paraclinical, and imaging features of COIVD-19.

## Background

In December 2019, the first case of unknown origin pneumonia was identified in Wuhan, the capital city of Hubei province. By Jan 7, 2020, Chinese scientists had isolated a novel virus belongs to coronaviruses family and classified it as a type of RNA virus[1].

Although primary zoonotic transmission of this virus, associated with a large seafood market (Huanan seafood market), has started the outbreak, person-to-person transmission of the virus started a pandemic involving 197 countries [2] [3].

The clinical outcomes of COVID-19 infection are various, including asymptomatic infection, mild upper respiratory tract illness, severe viral pneumonia, and even death. Patients are presented with various clinical manifestations such as fever, dyspnea, and cough[4].

Initially, some studies have observed particular imaging patterns on chest radiography and computed tomography in COVID-19 patients [5]. As our knowledge increased, recent studies claimed that the sensitivity of CT scan is higher compared to rRT-PCR in the diagnosis of SARS-Cov-2 infection[6].

Laboratory findings are essential in order to evaluate patients’ complications and triaging them [7]. Complete blood count as an easy and affordable test detects disorders such as leukopenia, anemia, and thrombocytopenia that are contributed to patients’ prognosis [8]. In response to inflammation induced by COVID-19 acute-phase reactants may increase or decrease [9]. These factors may contribute to the patient’s outcomes. This meta-analysis aims to measure the most common clinical, laboratory, and imaging findings among COVID-19 patients.

## Methods

### Protocol

In this study, we used a protocol based on the transparent reporting of systematic reviews and meta-analysis (PRISMA)(Table1).

**Table 1.**
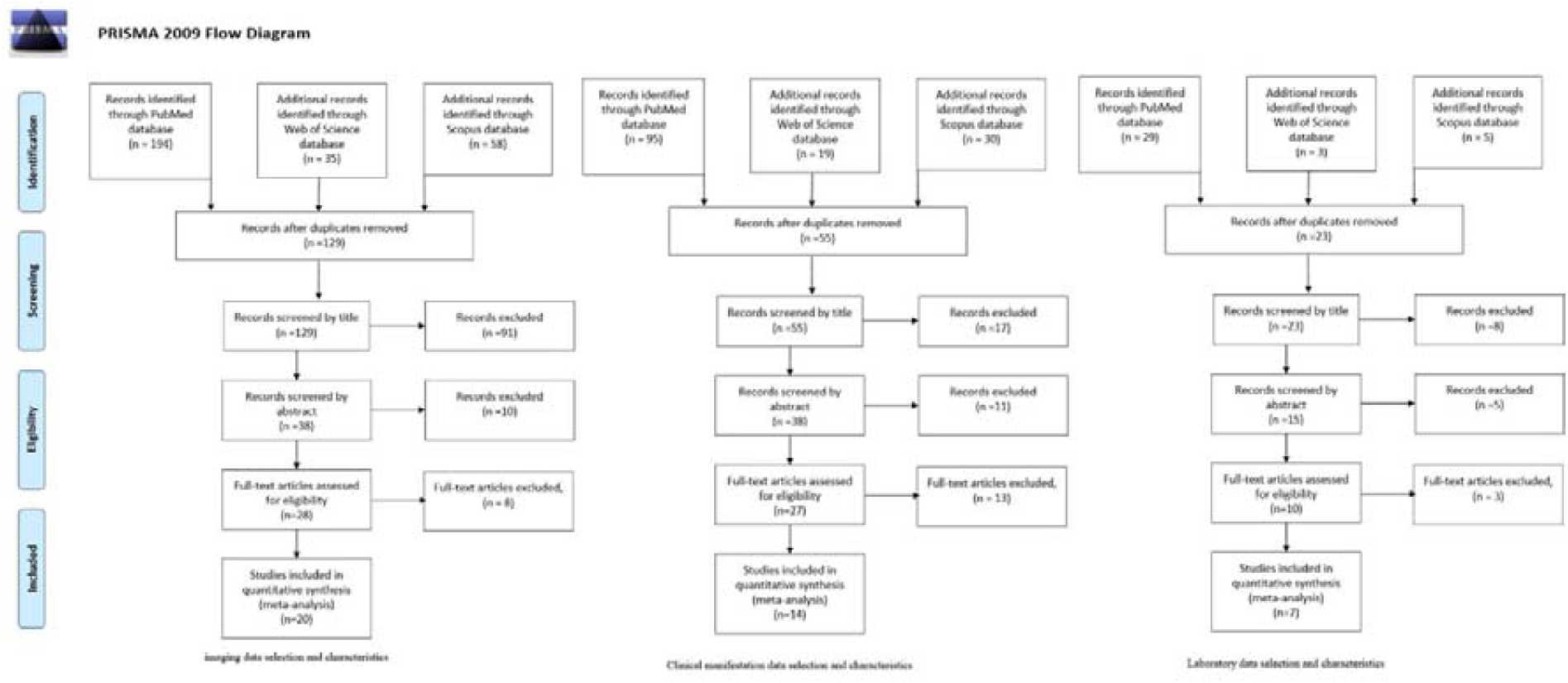
Study selection and characteristics

### Eligibility criteria

In this study, all included patients were confirmed using real-time reverse transcriptase-polymerase chain reaction (rRT-PCR). All searched articles were cohorts or case controls, and no language restrictions were conducted. All articles published before drafting the manuscript, where have been included. Review articles, opinion articles, and letters not presenting original data were excluded from the analysis.

### Information sources and search strategy

A systematic search was performed using Medline/PubMed, Scopus, and Web of Science based on following keywords: (“clinical manifestation” AND COVID-19) or (“clinical manifestation” AND 2019-nCoV) or (“clinical manifestation” AND COVID) or (“clinical manifestation” AND Corona) or (“clinical characteristics” AND COVID-19) or (“clinical characteristics” AND 2019-nCoV) or (“clinical characteristics” AND COVID) or (“clinical characteristics” AND corona) or (liver AND COVID-19) or (liver AND 2019-nCoV) or (liver AND COVID) or (liver AND Corona) or (“blood gas” AND COVID-19) or (“blood gas” AND 2019-nCoV) or (“blood gas” AND COVID) or (“blood gas” AND corona) or (COVID-19 AND radiography) or (2019-nCoV AND radiography) or (Corona AND radiography) or (COVID AND radiography) or (COVID-19 AND radiographic) or (2019-nCoV AND radiographic) or (Corona AND radiographic) or (COVID AND radiographic) or (COVID-19 AND CT) or (2019-nCoV AND CT) or (Corona AND CT) or (COVID AND CT) or (COVID-19 AND “computed tomography”) or (2019-nCoV AND “computed tomography”) or (Corona AND “computed tomography”) or (COVID AND “computed tomography”) or (CBC AND corona) or (CBC AND COVID) or (CBC AND COVID-19) or (CBC AND 2019-nCoV). systematic search was conducted prior to Mar-25 2020 and three independent researchers evaluated all papers.

### Study selection

In the initial search, we assessed title and abstract, followed by a full-text evaluation based on previously described inclusion and exclusion criteria. When two articles reported one patient’s characteristics, we merged all reported data and assumed as a single individual. Descriptive studies reporting clinical symptoms, laboratory, and radiological findings were used to perform a meta-analysis

### Data collection process and data items

Three independent researchers filled data extraction forms containing study type, Journal, publication date, sample size, age, gender, clinical characteristics, laboratory, and radiological findings Conflicts were resolved by another researcher

### Assessment of methodological quality and risk of bias

For quality assessment, we used the appraisal tool for Cross-Sectional Studies (AXIS). Publication bias was assessed with a funnel plot for the standard error by logit event rate, with no evidence of bias (figure1,2, and 3). A mixed-effect model was used to calculate the pooled estimate and 95% CI. Due to limitations in several studies, results were reported in the fixed-effect model. Given variable degrees of data heterogeneity, and the inherent heterogeneity in any systematic review studies published in the literature Heterogenicity indexes such as Q-value, I-squared, and Tau Squared are mentioned in table 4, 5, and 6. Also, figures 4, 5, and 6 depict Pool prevalence Frost Plots of each data (clinical, laboratory, and imaging).

**Figure 1.**
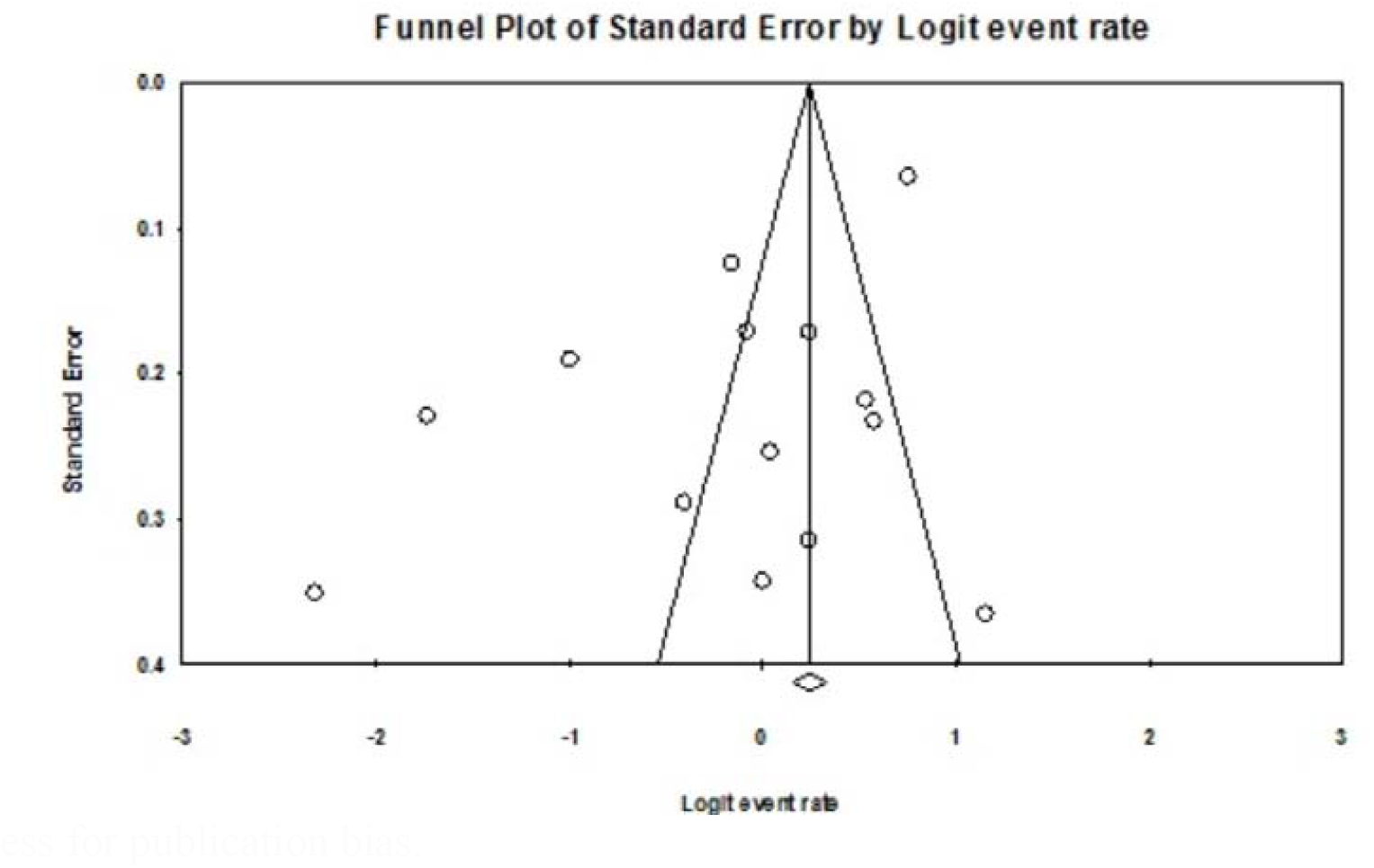
clinical characteristics funnel-plot for the Standard Error by Logit Event rate to assess for publication bias.

**Figure 2.**
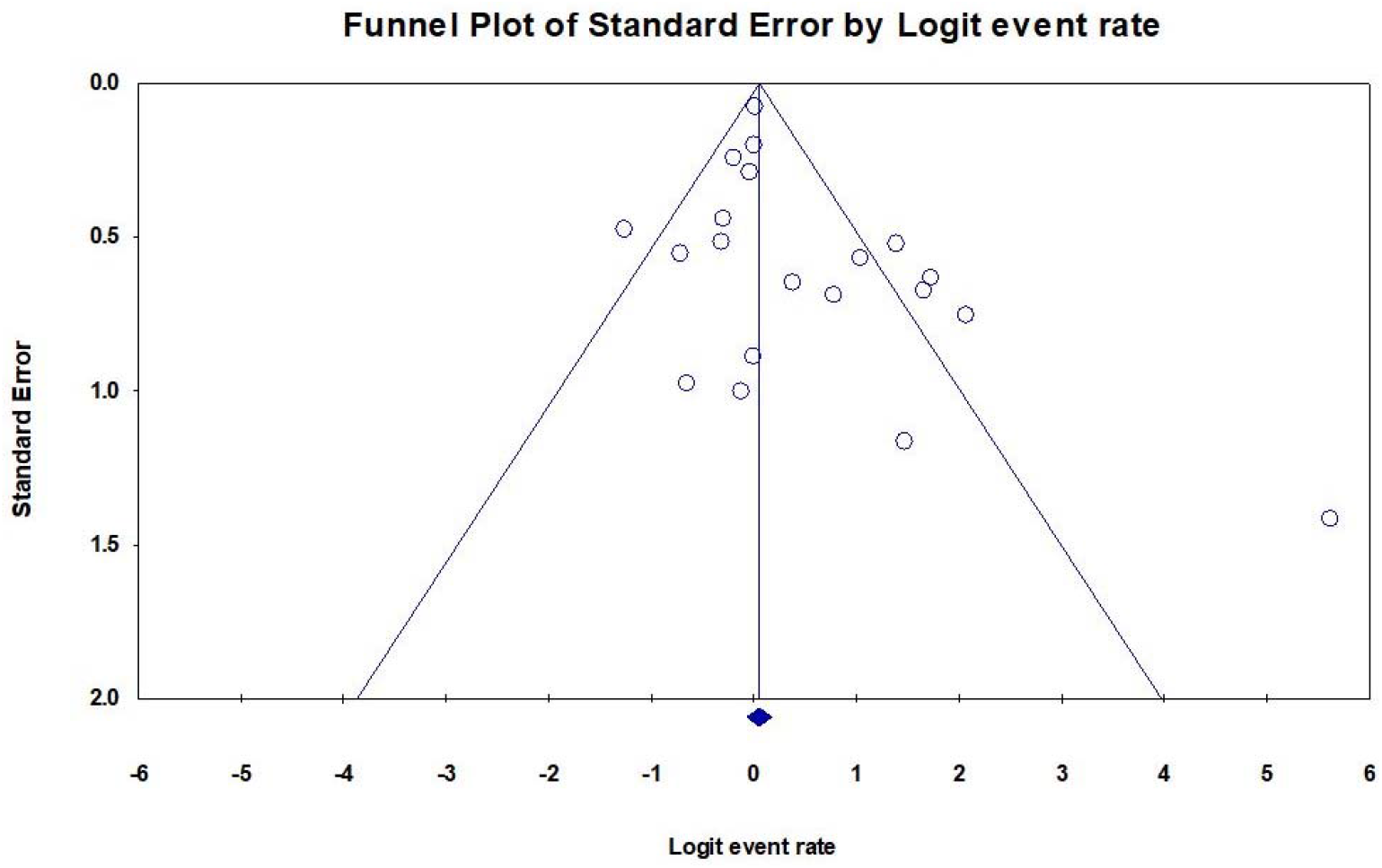
CT-scan funnel-plot for the Standard Error by Logit Event rate to assess for publication bias.

**Figure 3.**
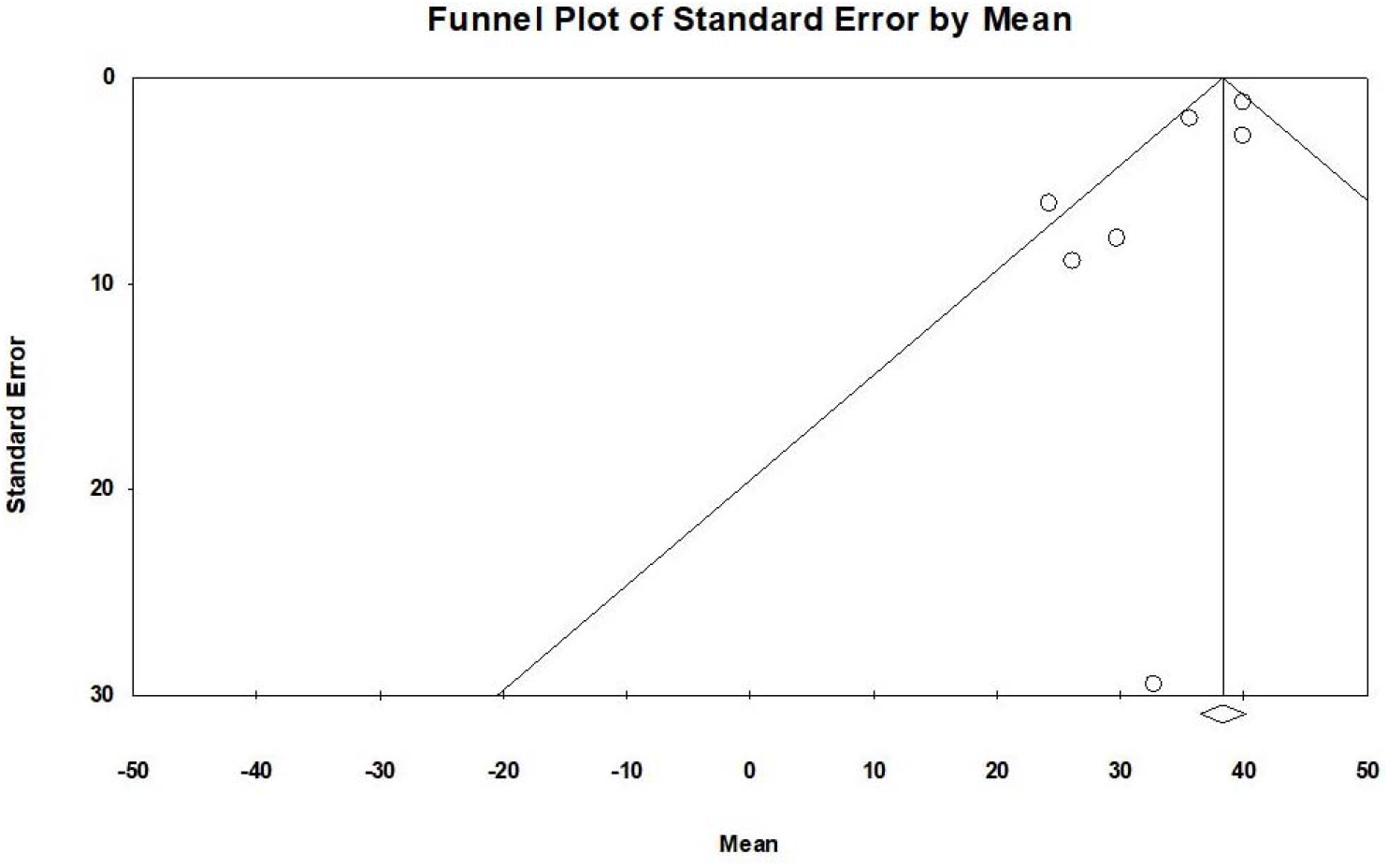
laboratory finding funnel-plot for the Standard Error by Logit Event rate to assess for publication bias.

**Figure 4.**
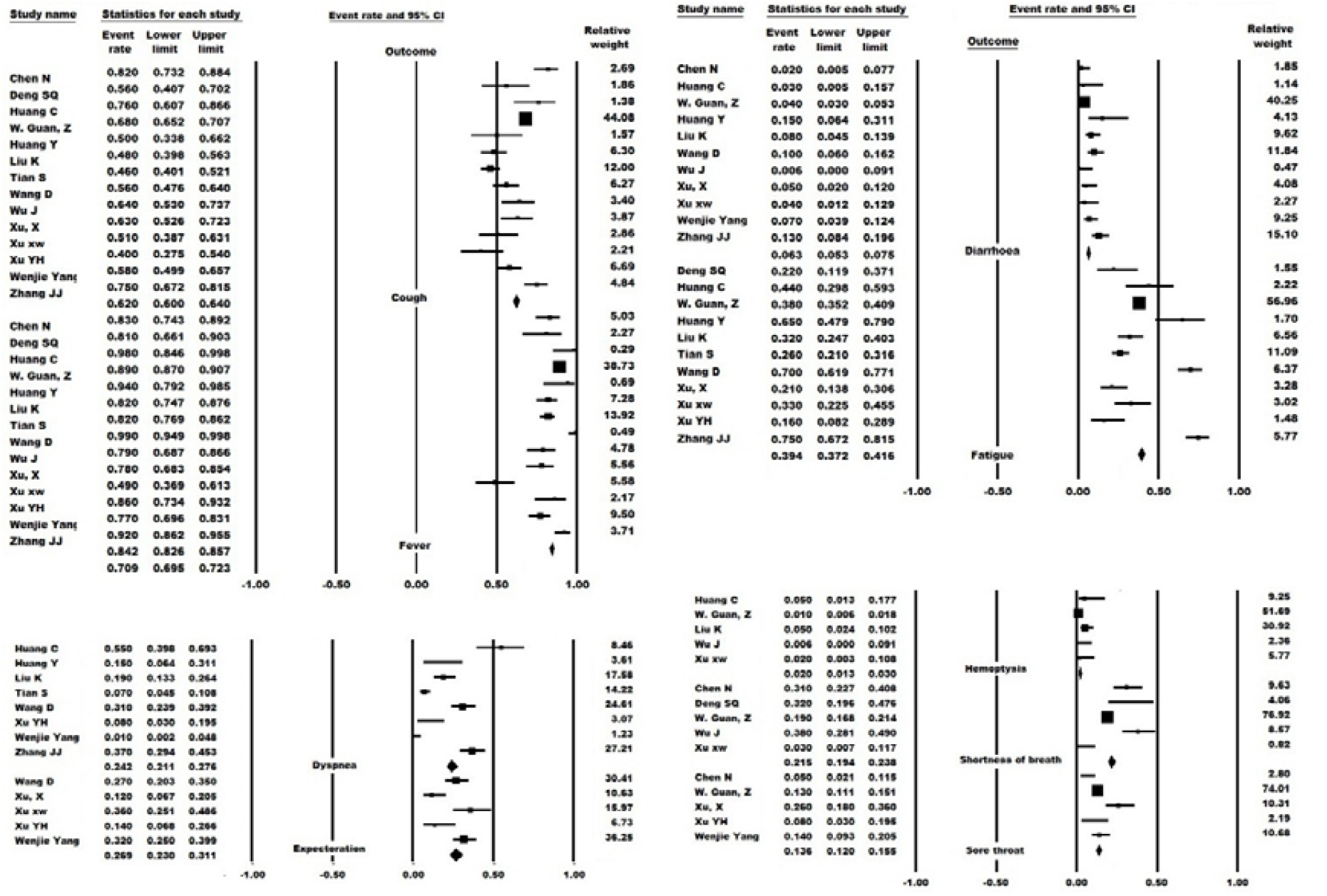
Pool prevalence forest plots of clinical manifestations

**Figure 5.**
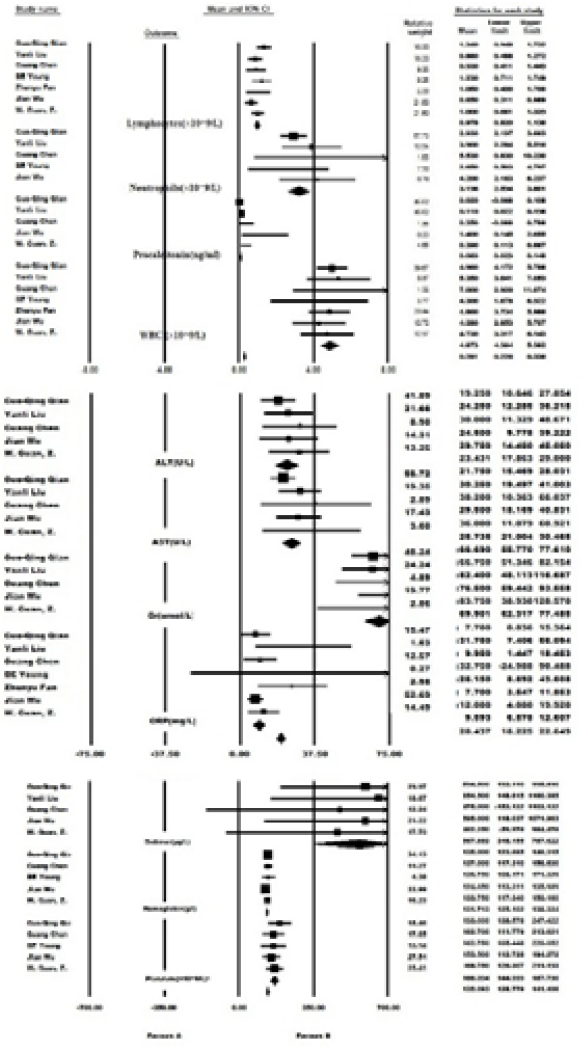
Pool prevalence for estplots of laboratory findings

**Figure 6.**
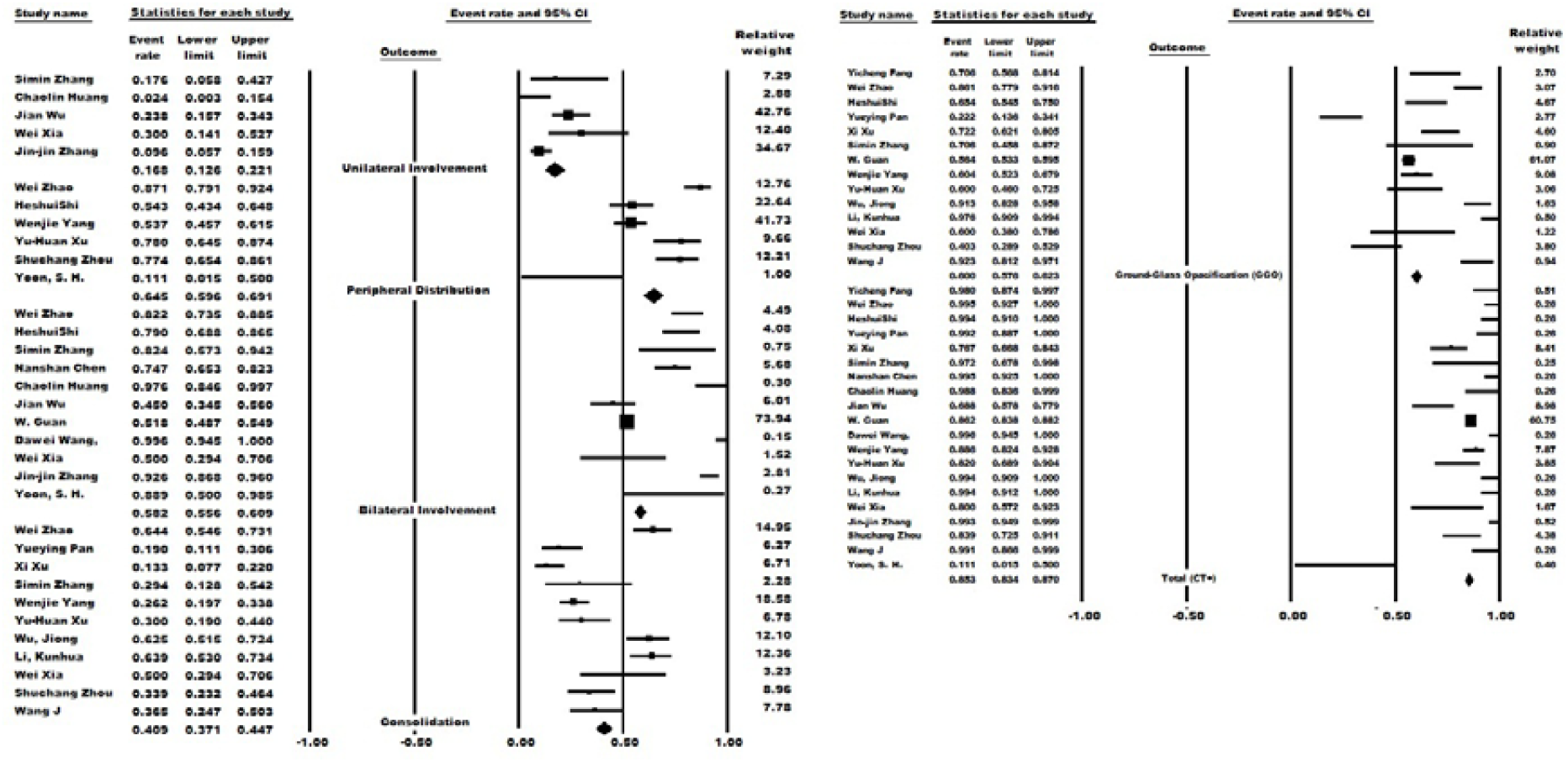
Pool prevalence forest plots of CT-scan findings

### Statistical approach

Effect size pooled estimate for imaging and clinical data, was measured based on Event rate, Logit event rate, and standard error.

Considering that the computational index of laboratory data was median in order to meta-analysis them in CMA v.2. software, we use the following formula

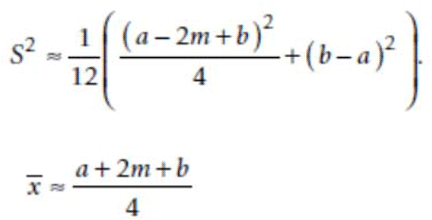

Estimating the mean and variance from the median

m = Median

a = The smallest value (minimum)

b = The largest value (maximum)

n = The size of the sample [10]

The meta-analysis was performed using Stata, the software Open Meta [Analyst], and Comprehensive Meta-Analysis Software (CMA) ve.2. Pooled estimate and their 95% confidence intervals (95% CIs) were used to summarize the weighted effect size for each study grouping variable.

## Results

### Study selection and characteristics

207 articles were included based on a search strategy, which, as previously described (Table1). The full text of 65 articles was evaluated after the title and abstract assessment. Twenty-four articles were excluded due to inadequate data. Finally, the meta-analysis was performed on 30 articles (three different Subjects). The article’s data Summary is reported in Table 2. Also, demographic characteristics and comorbidities of studies are demonstrated in Table 3.

**Table 2.** Characteristics of the included studies on COVID-19, 2020.

| Row | Author | Journal | Data | Country | Sample Size | Quality score<sup>a</sup> | Reference |
| --- | --- | --- | --- | --- | --- | --- | --- |
| **imag-** |  |  |  |  |  |  |  |
| **ing** |  |  |  |  |  |  |  |
| 1 | Yicheng Fang | Radiology | Feb-19 | china | 51 | 15 | (6) |
| 2 | Wei Zhao | American Journal of Roentgenology | Feb-18 | china | 101 | 16 | (11) |
| 3 | HeshuiShi | The Lancet | Feb-24 | china | 81 | 17 | (12) |
| 4 | Yueying Pan | European Radiology | Feb-13 | china | 63 | 14 | (13) |
| 5 | Xi Xu | European Journal of Nuclear Medicine and Molecular Imaging | Feb-28 | china | 90 | 15 | (14) |
| 6 | Simin Zhang | European Respiratory Journal | Mar-25 | china | 17 | 14 | (15) |
| 7 | Nanshan Chen | The Lancet | Jan-30 | china | 99 | 18 | (16) |
| 8 | Chaolin Huang | The Lancet | Feb-21 | china | 41 | 18 | (17) |
| 9 | Jian Wu | Clinical Infectious Diseases | Feb-29 | china | 80 | 12 | (18) |
| 10 | W. Guan | the New England Journal of Medicine | Feb-28 | china | 1099 | 19 | (19) |
| 11 | Dawei Wang, | JAMA | Feb-29 | china | 138 | 19 | (20) |
| 12 | Wenjie Yang | Journal of Infection | Feb-26 | china | 149 | 13 | (21) |
| 13 | Yu-Huan Xu | Journal of Infection | Feb-25 | china | 50 | 15 | (22) |
| 14 | Wu, Jiong | Investigative Radiology | Feb-29 | china | 80 | 17 | (23) |
| 15 | Li, Kunhua | Investigative Radiology | Feb-29 | china | 83 | 18 | (24) |
| 16 | Wei Xia | Pediatric Pulmonology | Mar-05 | china | 20 | 12 | (25) |
| 17 | Jin-jin Zhang | Allergy | Feb-19 | china | 140 | 12 | (26) |
| 18 | Shuchang Zhou | American Journal of Roentgenology | Feb-16 | china | 62 | 14 | (27) |
| 19 | Wang J | Journal of Zhejiang University | Feb-24 | china | 52 | 15 | (28) |
| 20 | Yoon, S. H. | Korean J Radiol | Apr-21 | china | 9 | 12 | (29) |
| **lab** |  |  |  |  |  |  |  |
| 21 | Guo-Qing Qian | An International Journal of Medicine | Feb-21 | china | 91 | 16 | (30) |
| 22 | Yanli Liu | MedRxiv |  | china | 109 | 15 | (31) |
| 23 | Guang Chen | MedRxiv |  | china | 21 | 12 | (32) |
| 24 | BE Young | Jama | Feb-24 | china | 18 | 17 | (33) |
| 25 | Zhenyu Fan | The Lancet | Mar-05 | china | 148 | 16 | (34) |
| 26 | Jian Wu | Clinical Infectious Diseases | Feb-29 | china | 80 | 12 | (18) |
| 27 | W. Guan, Z. | New England Journal of Medicine | Feb-28 | china | 1099 | 19 | (19) |
| **clinical** |  |  |  |  |  |  |  |
| 28 | Chen N | The Lancet |  | china | 99 | 18 | (16) |
| 29 | Deng SQ | Journal Clinical Medicine | Feb-14 | china | 41 | 15 | (35) |
| 30 | Huang C | The Lancet | Feb-21 | china | 41 | 18 | (17) |
| 31 | W. Guan, Z | the New England Journal of Medicine | Feb-19 | china | 1099 | 19 | (19) |
| 32 | Huang Y | Travel Medicine and Infectious Disease | Feb-24 | china | 34 | 13 | (36) |
| 33 | Liu K | Chinese medical journal | Feb-07 | china | 137 | 13 | (37) |
| 34 | Tian S | Journal of Infection | Feb-27 | china | 262 | 13 | (38) |
| 35 | Wang D | Jama | Feb-21 | china | 138 | 19 | (20) |
| 36 | Wu J | Clinical Infectious Diseases | Feb-29 | china | 80 | 12 | (18) |
| 37 | Xu, X | European Journal of Nuclear Medicine and Molecular Imaging | Feb-28 | china | 90 | 15 | (14) |
| 38 | Xu xw | BMJ : British Medical Journal | Feb-19 | china | 62 | 14 | (39) |
| 39 | Xu YH | Journal of Infection | Feb-25 | china | 50 | 14 | (22) |
| 40 | Wenjie Yang | Journal of Infection | Feb-26 | china | 149 | 14 | (21) |
| 41 | Zhang JJ | Allergy | Feb-19 | china | 140 | 15 | (26) |

**Table 3.**
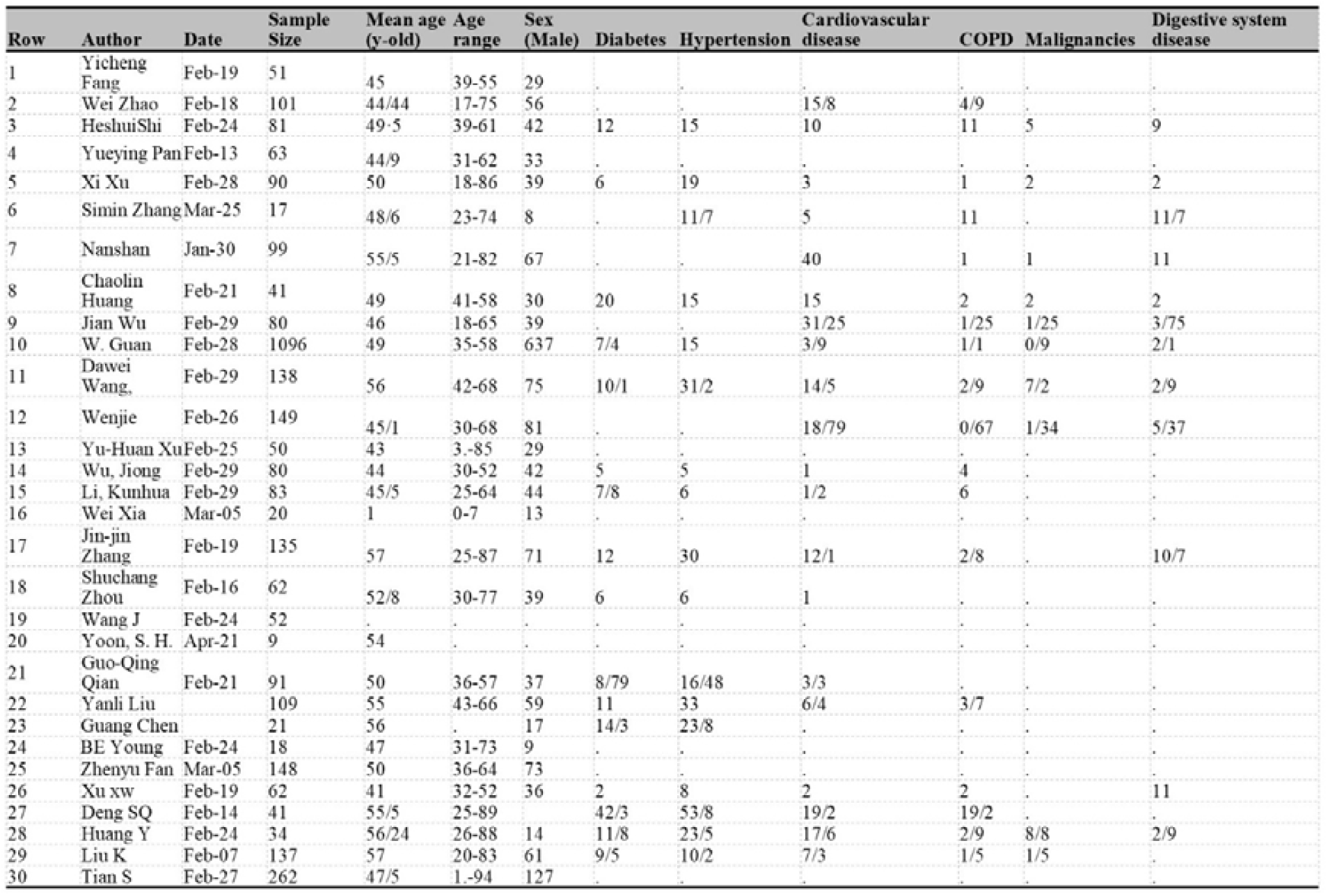
Demographical characteristics, and comorbidities of the study subjects

In this study, we evaluate 30 articles. all papers were from china, and 3420 individual’s data were evaluated. All studies were Cross-sectional, and 27 Variables were included.

### Clinical manifestations and laboratory findings

According to clinical manifestations, fever (84.2%, 95%CI 82.6-85.7), cough (62%, 95%CI 60-64), and fatigue (39.4%, 95%CI 37.2-41.6) are the most prevalent clinical symptoms among patients (Fig 4).

laboratory studies show increased level in following tests: CRP (9.59mg/l, 95%CI 6.58-12.61) with normal range of 0-3.0mg/l, D-dimer (567.89ng/ml, 95%CI 348.15-787.62) with normal range of 0-500ng/ml, LDH (255.96U/L, 95%CI 209.26-302.66) with normal range of 135-250U/L, and procalcitonin (0.09ng/ml 95%CI 0.03-0.15) which is normally less than 0.05ng/ml in healthy individual.

Also, the level of some laboratory factors are lower than normal; such as lymphocyte (0.97*10^9/L, 95%CI 0.03-0.15), Albumin (38.94g/L, 95%CI 37.5-40.83), and PaO2(73.77mmHg, 95%CI 52.8-94.74) concerning the normal range of 1-4*10^9/L, 40-55g/L, and 83-108mmHg respectively (Fig 5).

### CT-scan finding

Among all patients infected by COVID-19 (confirmed by RT-PCR), 85% had abnormalities in CT-scans. In most of them, the Bilateral pneumonia was dominant (58% 95CI 56-61), other image findings were: ground-glass opacities (GGO) (60%, 95%CI 58-62), Peripheral Distribution (64%, 95%CI 60-69), and Consolidation (41%, 95%CI 37-45) in those with CT-scan results (Fig 6).

## Discussion

From Dec-2019, more than 500,000 cases of new unknown origin pneumonia have been confirmed all over the world [40]. It was primarily known as 2019-nCov, then WHO decided to name this novel coronavirus “SARS-Cov2” [41]. COVID-19 is a severe condition that is compromising the health condition of people in all countries worldwide [42]. Identifying the various characteristics of this infection is vital for controlling the outbreak in different countries [43]. Clinical, laboratory, and image findings are essential to evaluate the different aspects of infection [44]. Different outcomes of COVID-19 (from asymptomatic infection to death) and Contagiousness of this virus, even in its incubation period [45], emphasize why discovering different characteristics are crucial in controlling this pandemic.

In this systematic review and meta-analysis, we describe the most common clinical data on COVID-19 confirmed cases that were published during the first months of the outbreak. We analyze 2422, RT-PCR confirmed patients, for different clinical manifestations. Our findings are robust due to the pooled results after combining all the studies’ data.

As expected from initial studies in China, COVID-19 patients presented predominantly with cough and fever, as well as headache, diarrhea, and fatigue, among other clinical features [46]. This was consistently found in many of the included studies [18, 47]. Fever frequency is similar in other β-CoV associated infections such as SARS and MERS, but studies showed that the cough frequency is higher in SARS and COVID-19 than MERS (< 50%) [48, 49]. In SARS and MERS, diarrhea is reported in about a quarter of patients, but our data shows that only 6 percent of COVID-19 patients present with diarrhea (Table 4). Our data also suggest that about 11 percent of patients are presented with headache as a symptom. Unlike SARS, which is well-characterized in the two-stage clinical course of the disease [50], COVID-19, still needs further definition to identify the Disease process.

**Table 4.** Meta-analysis of clinical manifestations (fixed-effects model).

| Group | Number<br>Studies | Point<br>estimate | 95% CI |  | Test of<br>null (2-Tail) |  |
| --- | --- | --- | --- | --- | --- | --- |
|  |  |  | Lower<br>limit | Upper<br>limit | Z-value | P-value |
| **Cough** | 14 | 0.620 | 0.600 | 0.640 | 11.445 | 0.000 |
| **Fever** | 14 | 0.842 | 0.826 | 0.857 | 27.883 | 0.000 |
| **Headache** | 12 | 0.116 | 0.103 | 0.131 | -29.802 | 0.000 |
| **Diarrhea** | 11 | 0.063 | 0.053 | 0.075 | -27.648 | 0.000 |
| **Fatigue** | 11 | 0.394 | 0.372 | 0.416 | -9.181 | 0.000 |
| **Dyspnea** | 8 | 0.242 | 0.211 | 0.276 | -12.503 | 0.000 |
| **Expectoration** | 5 | 0.269 | 0.230 | 0.311 | -9.472 | 0.000 |
| **Hemoptysis** | 5 | 0.020 | 0.013 | 0.030 | -17.905 | 0.000 |
| **Shortness of breath** | 5 | 0.215 | 0.194 | 0.238 | -19.180 | 0.000 |
| **Sore throat** | 5 | 0.136 | 0.120 | 0.155 | -23.906 | 0.000 |
| **Muscle ache** | 4 | 0.139 | 0.104 | 0.184 | -10.763 | 0.000 |

Studies on epidemiological features of COVID -19 showed that about 80 percent of patients are asymptomatic or are presented with mild manifestations [51, 52], but almost all of the patients included in our study had moderate to severe characteristics. It seems that fever and cough are the most common clinical features among moderate to severe patients (Table 4)

Studies show different laboratory abnormalities in COVID-19 infected patients, such as hypoalbuminemia or elevated inflammatory markers [53]. However, our data suggest that C-reactive protein is the most elevated factor among infected cases (Table 5). D-dimer, LDH, and procalcitonin are also elevated in patients, which confirmed that measuring inflammatory markers are essential to investigate new cases [54]. Also, seven studies showed lymphopenia and albuminuria as other common laboratory findings. Data from new studies suggest lymphocytopenia or an increase in WBC as prognostic factors in COVID-19 patients. Studies on the SARS outbreak in 2003 indicate that lymphopenia, leukopenia, and thrombocytopenia, elevated levels of LDH, alanine transaminase (ALT), AST, and creatine-kinase are most affected laboratory findings [55].

**Table 5.** A meta-analysis of laboratory outcomes (fixed-effects model)

| Group | Number<br>Studies | Point estimate<br>(Mean) | 95% CI |  | Standard<br>error | Variance | Z-value | P-value |
| --- | --- | --- | --- | --- | --- | --- | --- | --- |
|  |  |  | Lower limit | Upper limit |  |  |  |  |
| CRP (mg/L) | 7 | 9.59 | 6.58 | 12.61 | 1.54 | 2.37 | 6.24 | 0.000 |
| Lymphocytes($\times 10^9/L$) | 7 | 0.98 | 0.82 | 1.14 | 0.08 | 0.01 | 12.15 | 0.000 |
| WBC($\times 10^9/L$) | 7 | 4.87 | 4.36 | 5.38 | 0.26 | 0.07 | 18.76 | 0.000 |
| ALT(U/L) | 5 | 23.43 | 17.86 | 29.00 | 2.84 | 8.07 | 8.25 | 0.000 |
| AST(U/L) | 5 | 25.73 | 21.00 | 30.47 | 2.41 | 5.83 | 10.66 | 0.000 |
| Cr ($\mu\text{mol/L}$) | 5 | 69.90 | 62.32 | 77.48 | 3.87 | 14.97 | 18.06 | 0.000 |
| D-dimer($\mu\text{g/L}$) | 5 | 567.89 | 348.15 | 787.62 | 112.11 | 12568.89 | 5.07 | 0.000 |
| Hemoglobin(g/l) | 5 | 131.71 | 125.10 | 138.32 | 3.37 | 11.38 | 39.05 | 0.000 |
| LDH(U/L) | 5 | 255.96 | 209.26 | 302.66 | 23.83 | 567.76 | 10.74 | 0.000 |
| Neutrophils($\times 10^9/L$) | 5 | 3.20 | 2.59 | 3.80 | 0.31 | 0.09 | 10.38 | 0.000 |
| Platelets($\times 10^9/L$) | 5 | 166.39 | 145.00 | 187.79 | 10.92 | 119.17 | 15.24 | 0.000 |
| Procalcitonin (ng/ml) | 5 | 0.09 | 0.03 | 0.15 | 0.03 | 0.00 | 2.90 | 0.004 |

Nevertheless, not significantly seen in COVID. SARS-Cov-2 can affect the liver and other organs [56]. AST and ALT are normal in most cases, but impaired liver function tests are associated with poor prognosis and higher mortality rates [34, 57]. Coagulation function tests (such as INR) are affected in the prognosis of this infection [58]. Lymphopenia in COVID -19 infected patients suggest that this virus might act on lymphocytes (mainly T cells), but some studies suggest that B cells are also affected [59, 53].

Ct-scan is one of the most useful methods of respiratory disease diagnosis. CT scans high sensitivity and availability make it one of the most common tests for lung disease screening [60, 61]. In COVID-19 patients, different results could present in the early stages of infection [62, 63]; even some studies demonstrate that CT scans sensitivity is higher than rRT-PCR [6]. Our data shows that 92% of rRT-PCR confirmed cases had abnormal CT-scan results, which suggest CT-scan as a reliable method.

As seen in Table 6, CT-scan meta-analysis outcomes are performed in fixed and random effect analyses. Our meta-analysis on 15 studies showed that Ground-Glass Opacification (GGO) and peripheral distribution is seen on 60% of and 64% of patients, respectively. 58% of the patients had Bilateral Involvements, which is contributed to poor prognosis.

**Table 6.** Meta-analysis of CT-scan outcomes (random-effects model and fixed-effect model)

| Group | Number<br>Studies | Point<br>estimate | 95%CI |  | Test of<br>null (2-Tail) |  | Heterogeneity |  | P-value | I-squared |
| --- | --- | --- | --- | --- | --- | --- | --- | --- | --- | --- |
|  |  |  | Lower<br>limit | Upper<br>limit | Z-value | P-value | Q-value | df (Q) |  |  |
| Fixed effect analysis |  |  |  |  |  |  |  |  |  |  |
| Ground-Glass<br>Opacification (GGO) | 14 | 0.60 | 0.58 | 0.62 | 8.02 | 0.0000 | 139.79 | 13.00 | 0.0000 | 90.70 |
| Peripheral<br>Distribution | 6 | 0.64 | 0.60 | 0.69 | 5.62 | 0.0000 | 45.25 | 5.00 | 0.0000 | 88.95 |
| Bilateral Involvement | 11 | 0.58 | 0.56 | 0.61 | 6.04 | 0.0000 | 143.51 | 10.00 | 0.0000 | 93.03 |
| Consolidation | 11 | 0.41 | 0.37 | 0.45 | -4.59 | 0.0000 | 105.98 | 10.00 | 0.0000 | 90.56 |
| Total (CT+) | 20 | 0.85 | 0.83 | 0.86 | 24.34 | 0 |  |  |  |  |
| Random effects analysis |  |  |  |  |  |  |  |  |  |  |
| Ground-Glass<br>Opacification (GGO) | 14 | 0.70 | 0.60 | 0.78 | 3.89 | 0.0001 |  |  |  |  |
| Peripheral<br>Distribution | 6 | 0.67 | 0.50 | 0.80 | 1.96 | 0.0506 |  |  |  |  |
| 4)Bilateral<br>Involvement | 11 | 0.79 | 0.67 | 0.88 | 4.08 | 0.0000 |  |  |  |  |
| Consolidation | 11 | 0.38 | 0.26 | 0.51 | -1.85 | 0.0650 |  |  |  |  |
| Total (CT+) | 20 | 0.92 | 0.87 | 0.95 | 9.29 |  |  |  |  |  |

CT-scan is useful in monitoring the treatment, and it is crucial in classifying patients and identifying who should be treated with aggressive treatments [11]. Other findings such as consolidation or reverse halo or atoll sign are reported in some studies [64], which were not included in our analysis.

## Limitations

This review has several limitations. Few studies are available on COVID-19, and most of them are from China. Many countries such as Italy, the United States, and Iran reported several new SARS-Cov-2 positive patients, but data about clinical characteristics or laboratory findings are limited. By publishing more studies worldwide, researchers are going to get a more comprehensive understanding of COVID19. Patients, detailed information, especially in clinical outcomes, was unavailable in most studies at the time of analysis.

## Conclusions

COVID-19 presents in the majority of cases with fever and cough. Laboratory findings such as elevated inflammatory markers can help to diagnose. Other factors, such as AST, ALT, or INR, are associated with the prognosis of the patients. 92% of the RT-PCR confirmed that patients have abnormalities in CT-scan most frequently GGO. Additional research with higher sample sizes is needed in order to describe the patient’s characteristics more precisely.

## Data Availability

-

## Abbreviations

COVID-19: Corona Virus Disease of 2019
rRT-PCR: real-time Reverse Transcription–Polymerase Chain Reaction
CT-scan: Computed Tomography scan
CI: Confidence Intervals
CRP: C-Reactive Protein
RNA: Ribonucleic Acid
SARS-CoV-2: Severe Acute Respiratory Syndrome Corona Virus #2
PRISMA: protocol based on transparent reporting of systematic reviews and meta-analysis
CBC: Complete Blood Count
CMA: comprehensive meta-analysis
GGO: ground-glass opacities
β-CoV: beta coronavirus
MERS: Middle Eastern Respiratory Syndrome
SARS: Severe Acute Respiratory Syndrome
LDH: Lactate Dehydrogenase
WBC: White Blood Cell
ALT: Alanine Transaminase
AST: Aspartate Aminotransferase
INR: International Normalized Ratio.

## Authors contribution

HH: Managed the group and participated in preparing the manuscript. SS: participated in meta-analysis. SM & NT: participated in the systematic search substantially drafted the manuscript. PM revised the manuscript critically. All authors read and approved the final version of the manuscript.

